# Peripheral blood mononuclear cell transcriptomic trajectories reveal dynamic regulation of inflammatory actors in delirium

**DOI:** 10.1101/2025.02.14.25322163

**Authors:** Sara C. LaHue, Naoki Takegami, Rubinee Simmasalam, Abiya Baqai, Elena Munoz, Anya Sikri, Thibault du Buisson de Courson, Nilika S. Singhal, Walter Eckalbar, Charles R. Langelier, Carolyn M. Hendrickson, Carolyn S. Calfee, David J. Erle, Matthew F. Krummel, Prescott G. Woodruff, Tomiko Oskotsky, Marina Sirota, Adam Ferguson, Vanja C. Douglas, John C. Newman, Samuel J. Pleasure, Michael R. Wilson, COMET consortium, Neel S. Singhal

## Abstract

Delirium is a neurologic syndrome characterized by inattention and cognitive impairment frequently encountered in the medically ill. Peripheral inflammation is a key trigger of delirium, but the patient-specific immune responses associated with delirium development and resolution are unknown. This retrospective cohort study of prospectively collected biospecimens examines RNA sequencing from peripheral blood mononuclear cells of adults hospitalized for COVID-19 to better understand patient-specific factors associated with delirium (n = 64). Longitudinal transcriptomic analyses highlight persistent immune dysregulation in delirium, marked by increasing expression trajectories of genes linked to innate immune pathways, including complement activation, cytokine production, and monocyte/macrophage recruitment. Genes involved adaptive immunity showed a declining trajectory over time in patients with delirium. Although corticosteroid treatment suppressed some aspects of immune hyperactivation, aberrant responses contributing to delirium were exacerbated. Delirium resolution was characterized by normalization of key transcripts such as *CCL2* and innate immune markers. Novel associations with delirium were found in genes related to stress granule assembly and *DUSP2* and *KLF10*, which mediate T-cell responses. These findings provide insights into the peripheral immune responses accompanying delirium and their modulation by corticosteroids. Future trials targeting aberrant inflammatory responses may mitigate the severe outcomes associated with delirium due to COVID19.

## INTRODUCTION

Among patients requiring hospitalization for hypoxia related to severe acute respiratory syndrome coronavirus (SARS-CoV)-2 infection, the development of delirium is associated with worse outcomes, especially in older patients ^1–3^. Delirium is a common complication of hospitalization in adults and is characterized by an acute change in cognition and fluctuating attention. Over 30% of adults admitted to the hospital for medical conditions or surgery develop delirium during their hospitalization, and up to 70% of patients admitted to the intensive care unit (ICU) develop delirium ^4–8^. Patients with delirium experience longer hospitalizations and greater morbidity and mortality than those without delirium. Major risk factors for delirium include advanced age, dementia, the use of sedating medications, and acute infection ^9^. Identification of patients most likely to benefit from intensive, multicomponent non-pharmacologic delirium prevention programs may further help reduce complications and cost of care ^10,11^. The identification of delirium relies upon clinical observations and bedside assessments, with no reliable diagnostic imaging- or blood-based tests. The heterogenous nature of delirium complicates development of a single blood-based biomarker; however, recent studies have pointed to plausible common pathophysiological mechanisms including exaggerated immunologic responses and increased blood-brain barrier permeability leading to altered neurotransmission ^12–14^.

Severe COVID-19 is associated with excessive inflammatory responses, coagulopathy, and endothelial dysfunction, all factors that may contribute to the pathophysiology of delirium. As part of the University of California San Francisco (UCSF) COMET (COVID-19 Multi-phenotyping for Effective Therapies) and the NIAID IMPACC (Immunophenotyping Assessment in a COVID-19 Cohort) studies, serial biospecimens were prospectively collected, providing a unique opportunity to evaluate transcriptomic responses related to delirium over time. To further our understanding of the underlying biological mechanisms associated with the development of delirium in hospitalized patients with COVID-19, we analyzed transcriptomic data from peripheral blood mononuclear cells (PBMCs) isolated from hospitalized patients with COVID-19, with and without delirium, at admission and/or hospital days 4, 7, 14, 21, or 28. These rich clinical and biological data allowed—for the first time—longitudinal analyses elucidating the relationship between peripheral inflammation, timing of delirium (e.g., onset, during, and resolution), and corticosteroid treatment which greatly add to our understanding of the pathophysiology of delirium.

## MATERIALS AND METHODS

### Cohort selection

Adult patients admitted to UCSF Hospitals or Zuckerberg San Francisco General Hospital (ZSFG) with known or presumptive COVID-19 were screened within 3 days of hospitalization for inclusion in COMET/IMPACC. Patients, or a designated surrogate, provided informed consent to participate in the study in accordance with protocols approved by the Institutional Review Board of UCSF and ZSFG (#20-30497). This study includes patients enrolled between April 8, 2020 and May 1, 2021 in the COMET (COVID-19 Multi-immunophenotyping projects for Effective Therapies; https://www.comet-study.org/) and/or IMPACC studies at UCSF. IMPACC and COMET are prospective studies with aligned protocols that aim to describe the relationship between specific immunologic assessments and the clinical courses of COVID-19 in hospitalized patients. For inpatients, clinical data were abstracted from the electronic medical record into standardized case report forms. Patients were included in this analysis only if plasma was collected within 24 hours of patient admission or at least 3 serial samples were collected during the hospitalization. All participants are identified with a random identification specific for the COMET study after removing variables that could result in patient reidentification.

Patients were defined as having prevalent delirium if at any time during hospitalization there was documentation of a positive CAM-ICU (for patients in the ICU at UCSF or ZSFG) or NuDESC ≥2 (for patients hospitalized in the medical wards at UCSF) or if delirium was listed an active problem in the daily primary team clinical note (for patients hospitalized in the medical wards at ZSFG). Both CAM-ICU and NuDESC were prospectively assessed by trained nursing staff during each nursing shift ^32^. Incident delirium was defined as the documentation of delirium on nursing-led delirium screening assessments or primary team notes in patients initially presenting without delirium. Patients presenting without delirium were defined based on a negative first CAM-ICU or NuDESC <2 and absence of documentation of delirium or encephalopathy in admission notes. Delirium resolution was defined based on CAM-ICU remaining negative or NuDESC remaining <2 for the remainder of the hospitalization after previously having been positive, or documentation of resolution of delirium in the daily primary team note. The number of days of delirium (NDD) was defined as the number of days from delirium onset to persistent delirium resolution or discharge/death.

### Biospecimen collection

Peripheral blood was collected at admission and serially throughout the hospitalization (at days 4, 7, 14, and 28, unless discharge/death occurred prior to day 28) in sterile vacutainers after patient consent and transported on ice for processing. Blood plasma was isolated by centrifuging samples at 1000*g* for 10 min at 4°C, followed by density gradient centrifugation with SepMate PBMC isolation tubes (STEMCELL Technologies). Samples were then aliquoted and stored at -80°C until analysis.

### RNA isolation and sequencing

RNA was extracted from PBMCs using the Quick RNA MagBead kit (Zymo Research) on a KingFisher Flex system (Thermofisher Scientific) according to the company’s protocol. RNA integrity was measured with the Fragment Analyzer (Agilent) and library generation was continued when integrity was at least 6. Total RNA-sequencing libraries were depleted from ribosomal and hemoglobin RNAs, and generated using FastSelect (Qiagen) and Universal Plus mRNA-seq with Nu Quant (Tecan) reagents. Pooled libraries were PE100 sequenced on an HiSeq4000 or PE150 sequenced on an Illumina NovaSeq 6000 S4 flow cell at the Chan Zuckerberg Biohub SF.

### Bioinformatic and statistical analysis

Data analysis was performed in R using the statistical packages that are specifically mentioned below as well as the packages tidyverse, dplyr, ggplot2, EnhancedVolcano, volcano3D ^33–38^. The raw reads of the fastq files were tested for quality control using the FastQC software and were then aligned to the human reference genome (hg38 from the University of California, Santa Cruz) to summarize the RNA counts ^39^. DESeq2 was used for normalization and differential expression analysis of RNA counts ^40–42^. Differentially expressed genes were used as the input for gene ontology (GO) analysis with Enrchr-KG using GO: Biological Pathways ^43^. The normalized expression matrix generated was used as a template for WGCNA ^44^. CIBERSORTX (cibersortx.stanford.edu) analysis was applied to the gene expression matricies to impute immune cell fractions based on a signature matrix derived from single cell RNA sequencing of PBMCs classifying expression patterns into 17 cell types ^45^. Chi-Square tests were used to assess for statistical significance in the observed frequencies of categorical clinical data using Graphpad Prism 10 (La Jolla, CA).

## RESULTS

### Cohort information

The COMET cohort contained 64 subjects with at least 3 serial samples which were collected at hospital admission and/or hospital days 4, 7, 10, 14, 21, or 28 (Fig. 1A). Delirium was classified based on positive screening on Confusion Assessment Method (CAM) for the ICU (CAM-ICU), Nursing Delirium Scale Score (Nu-DESC) ≥2 for patients not in the ICU, and clinical team notes denoting delirium as an active problem. Clinical characteristics and outcomes between patients with delirium (n = 25) and those without delirium throughout the hospitalization (n = 39) are shown in Table 1. As expected, patients with delirium presented with more severe disease as indicated by sequential organ failure assessment (SOFA) and WHO COVID severity scores. In addition, length of hospitalization was also generally longer in patients developing delirium. However, a majority of patients in the cohort received ICU care and similar disease-specific treatments. Subsequent analyses comparing patients with prevalent delirium and no delirium are presented with adjustment for SOFA and WHO COVID severity classification scores by regression analysis. Additional information regarding delirium subjects, corticosteroid treatment, and onset/resolution of delirium is shown in Supplementary Figure 1.

**Fig 1:**
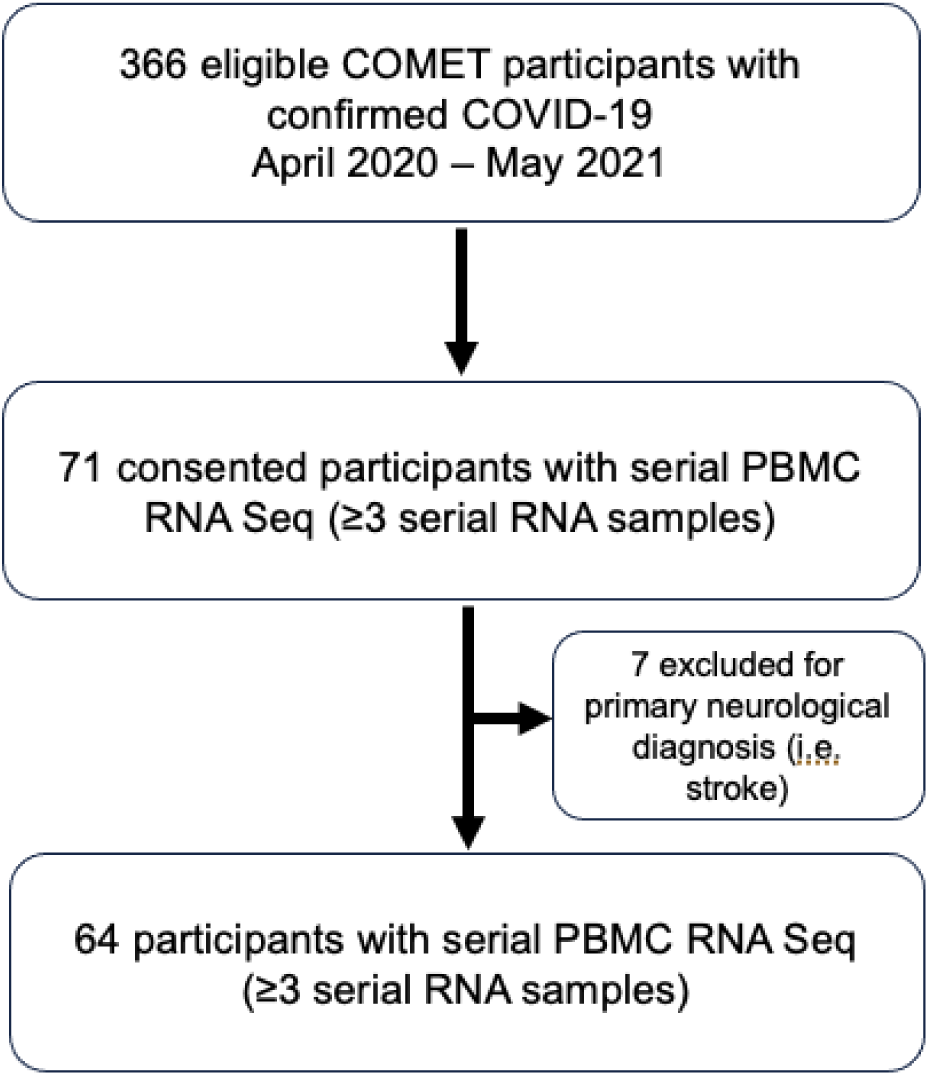
Flowchart of the COMET cohort used for peripheral blood mononuclear cell (PBMC) RNA-sequencing analyses. Patients were classified as having delirium based on positive screening results from nurse administered assessments (Confusion Assessment Method for Intensive Care Unit or the Nursing Delirium Screening Scale) and/or identification of active delirium in physician notes.

**Table 1.**
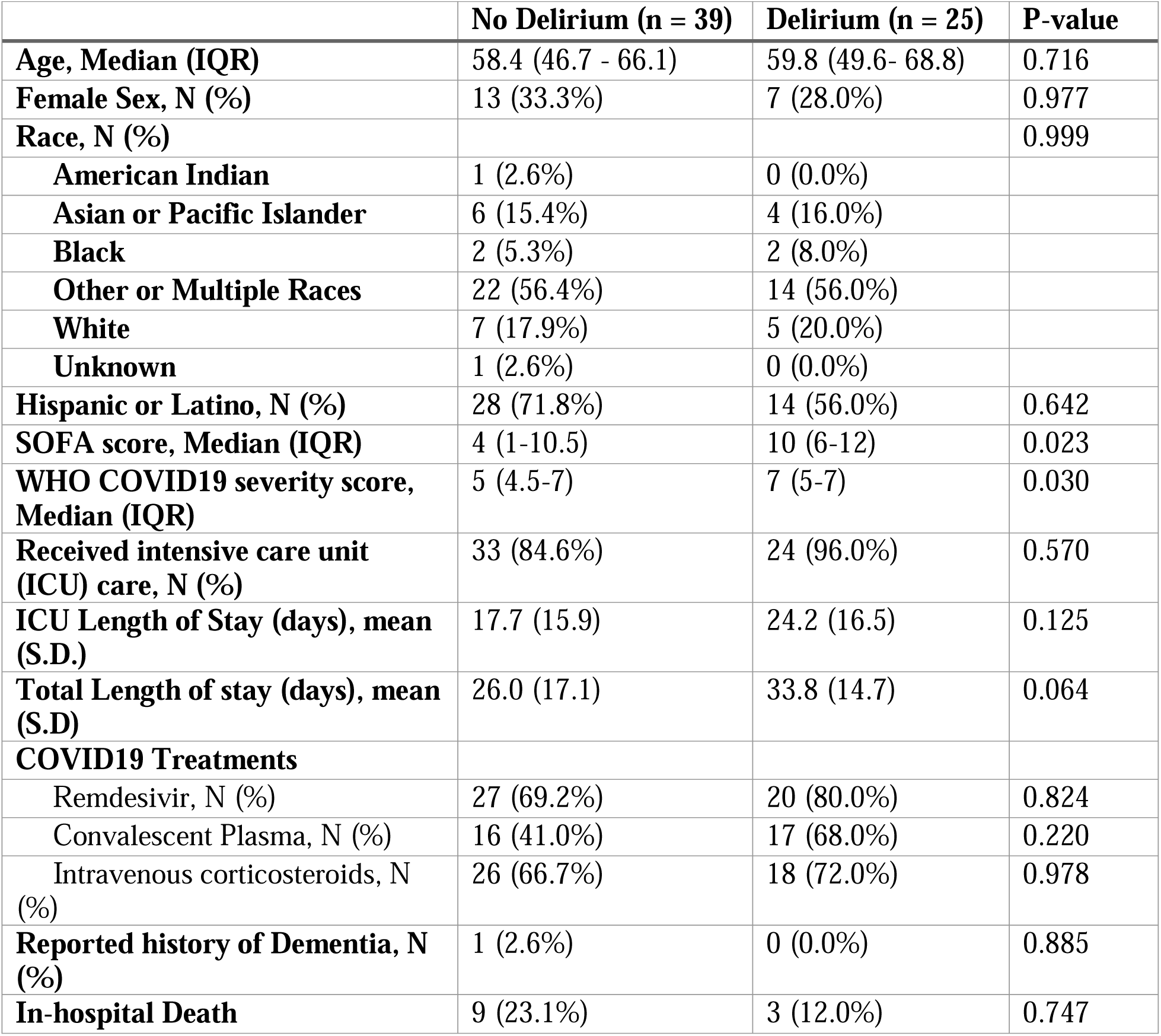
Patient demographics, treatments, and outcomes.

### Longitudinal transcriptomic signatures of delirium

A unique feature of our study, compared to prior transcriptomic studies in delirium, is the availability of serial sequencing to capture the transcriptomic response in those with prevalent delirium compared with those who did not develop delirium. Thus, analyses were performed to compare the transcriptomic trajectory over time in patients with prevalent delirium (n = 25) versus patients negative for delirium for the duration of hospitalization (n = 39). In comparing transcriptomic trajectories over time of delirium patients to patients that did not develop delirium we hope to identify key immune response signatures characteristic of delirium and generate candidate therapeutic pathways. We included only patients that were hospitalized for at least one week and with at least 3 serial samples to provide a robust comparison (Fig. 2A). We used a general linear mixed effects model (glmmSeq) adjusted for SOFA and WHO COVID severity scores to further dissect these longitudinal molecular signatures at the individual gene level. Unlike standard RNA sequencing analysis workflows, glmmSeq can fit negative binomial mixed models to allow for gene expression comparisons over time. Using glmmSeq for the longitudinal analysis, 547 genes were significantly up- or downregulated in delirium while 357 genes were differentially expressed in delirium over time based on the significance (FDR < 0.05) of the interaction term *time x diagnosis* (Supplementary Data File) and adjusted for disease severity. To further investigate pathway modulation in delirium, genes identified in the adjusted longitudinal mixed-effects model analysis were analyzed for gene ontology (GO): Biological Pathway (BP) enrichment. GO: BP analysis for genes significantly associated with the *time x diagnosis* include stress granule assembly, immunoglobulin production, lymphocyte mediated immunity, adaptive immune response, and angiogenesis genes among significantly enriched pathways (Fig. 2B). Interestingly, the GO:BP Learning includes genes which are enriched in the brain, but also well known to be expressed in PBMCs such as neurotensin receptor type 1, sodium-dependent serotonin transporter, neurabin-2, and synaptotagmin-11. Genes for key innate immune and inflammatory mediators demonstrated divergent trajectories over time with initially lower expression levels which increased over time in patients developing delirium compared to controls of C-C motif chemokine ligand (*CCL*)-2, *SLC6A19*, and *C1QA* (Fig. 2C). Similarly, T cell-interacting, activating receptor on myeloid cells (*TARM*)-1, plasma protease C1 inhibitor (*SERPING1*), and complement component receptor-1-like (*CR1L*), which are associated with potentially maladaptive T-cell and complement function also demonstrated persistently elevated expression delirium patients over time (Fig. 2C and Supplementary Fig. 2). Interestingly, tumor necrosis factor (*TNF*)-α, was also persistently elevated in delirium patients, but demonstrated a similar trajectory of decreased expression over time (Supplementary Fig. 2). Delirium was also associated with elevated expression of T-cell activating and chemotaxis-related genes in the initial days of hospitalization followed by marked decreases in expression including C-X-C chemokine receptor (*CXCR*)-3, interleukin-18 receptor (*IL18R*)-1, nuclear factor of activated T cells (*NFAT*)-c2, and *CD28* (Fig. 2C and Supplementary Fig. 2). Some genes involved in resolution of immune responses such as prostaglandin D2 receptor (*PTGDR*), *CCR5*, and *CD160* demonstrated increases over time in control patients, but were downregulated in patients with delirium. Interestingly, stress granule-associated genes, ataxin (*ATXN*)-2, cytoplasmic dynein 1 heavy chain (*DYNC1H*)-1, and proline rich coiled-coil (*PRCC*)-2C were significantly down-regulated over time in patients developing delirium which may also contribute to altered immune cell function. In summary, serial analysis of individual patient transcriptomic trajectories supports the hypothesis that an aberrant immune response characterized by initially exuberant innate immune activation and persistently elevated inflammatory mediators distinguish patients with delirium.

**Fig 2:**
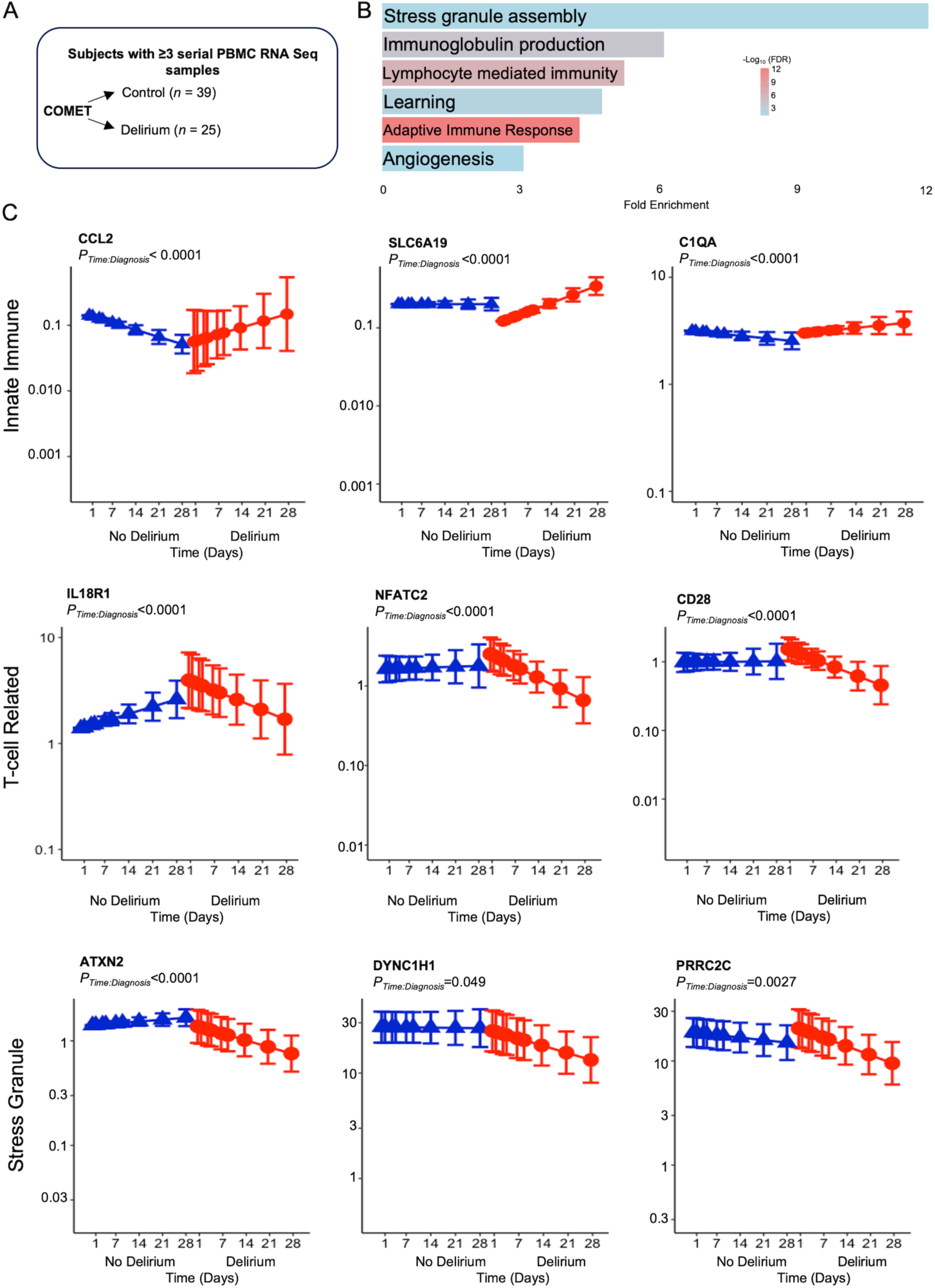
Mixed effects model of serial PBMC sequencing. **(A)** Schematic depicting cohort encompassed by generalized linear mixed-effects modeling (GLMM). **(B)** Gene ontology (GO): Biological pathway (BP) analysis of significantly differentially regulated RNAs in delirium patients over time. GO analysis of genes significant for *diagnosis x time* interaction in GLMM reveals significant regulation of immune-related pathways as well as those regulating stress granule assembly, and angiogenesis. **(C)** Plots for selected genes with colored points showing regression line of fitted mixed-effects model, with error bars showing 95% CIs (fixed effects).

We next examined how the transcriptomic response in patients with delirium varied as a function of time prior to, during, and after delirium resolution (n=25; Fig. 3A). We identified 2879 differentially expressed genes among sequencing before, during, and following delirium (FC > 0, FDR <0.01; Fig. 3B-D and Supplementary Fig. 3). Out of the genes differentially expressed in this analysis, 101 were specific to days where patients were positive for delirium, and 102 also overlapped with the *delirium x time* interaction in the severity-adjusted analysis above (out of 357). Key immune-related genes that were aberrantly low initially such as *CCL2*, *PTGDR*, *FCER1A*, and leukocyte immunoglobulin like receptor (*LILR*)-*A4*, increased with the resolution of delirium. Similarly, the resolution of delirium was associated with down-regulation of genes upregulated during delirium such as innate immune (*CR1L*, *SERPING1*, *ISG15*, *S100A8*, and *S100A9*) and inflammatory markers (*NFKBIA, IL18R1*, and immunoglobulin genes; Fig. 3D). These results show aberrant immune responses present at admission in patients that go on to develop delirium also resolve with delirium resolution.

**Fig. 3.**
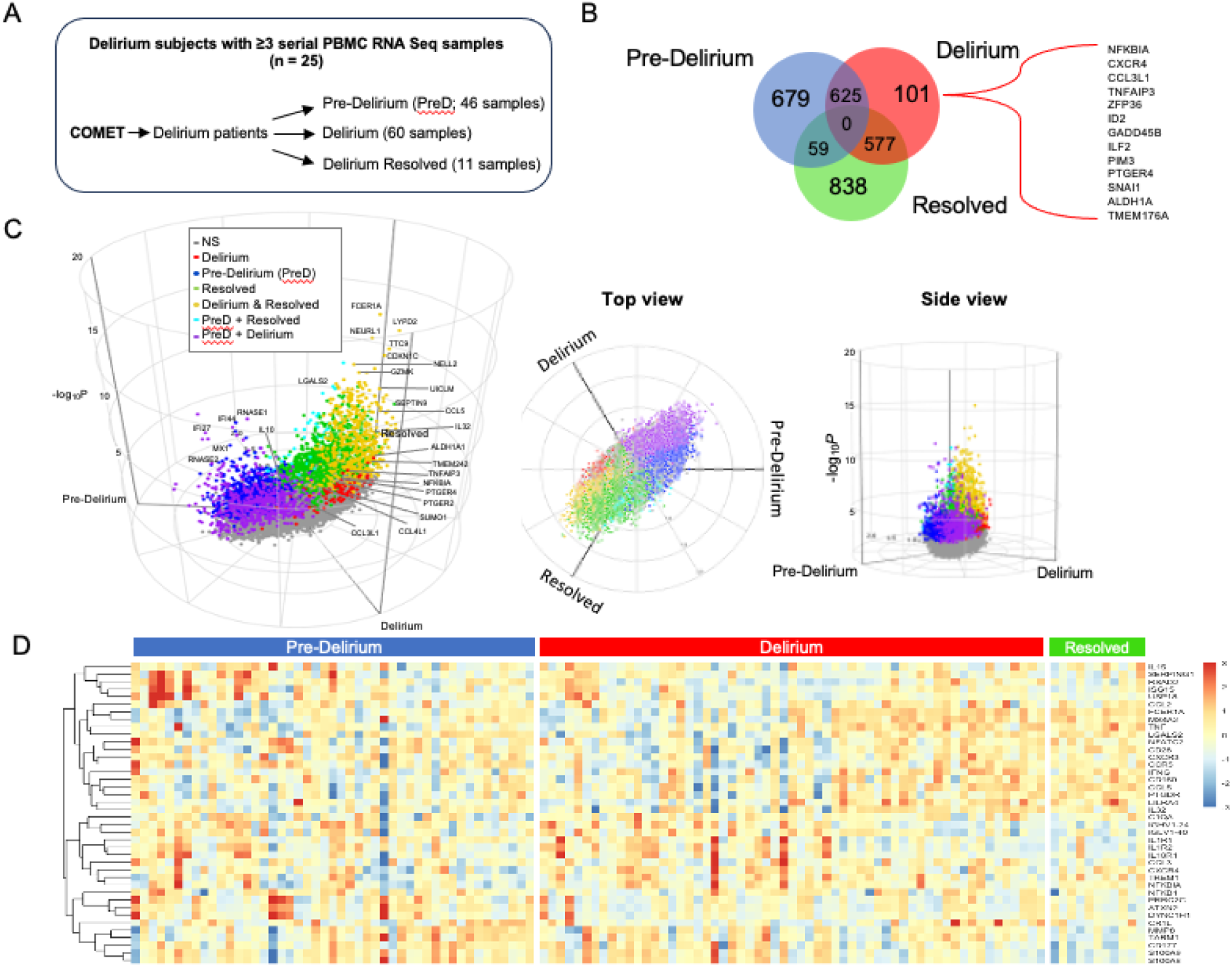
PBMC gene expression of delirium and delirium resolution. **(A)** Schematic of patients and samples included in the analysis. **(B)** Venn diagram demonstrating overlapping genes differentially regulated in samples prior to delirium (pre-delirium), during delirium, and the resolution of delirium. The highlighted gene list includes those also significant in the mixed-effects model analysis in Fig. 2. **(C)** 3D volcano plot demonstrating differentially expressed genes in patients prior to delirium (PreD), during delirium, and after delirium resolution. **(D)** Heat map demonstrating individual patient relative expression levels of selected significantly regulated genes in patients with delirium prior to, during, and after the resolution of delirium (FDR <0.01).

### Earlier onset of delirium is distinguished by innate immune activation

Mechanisms underlying the development of delirium at later time points (i.e. after days or weeks of illness, treatments, sedation, and sleep disturbances) may be fundamentally different than those contributing to early delirium when acute illness severity is high. When stratified according to early (<7 days, n = 11) versus late (≥7 days, n = 14) onset of delirium (Fig. 4A), mixed-effects modeling demonstrated contrasting changes in gene expression over time. Early compared to late onset of delirium had a significant effect on 533 genes, with 316 showing significant (FDR < 0.05) differential expression change over time. Patients with late onset delirium demonstrated increasing trajectories in *NFKBIA*, lymphocyte-specific protein tyrosine kinase (*LCK*), *IL32*, and *CXCR4*. While early onset delirium was associated with greater elevations in innate immune markers (*MMP8*, *S100P*), *CD177* (a neutrophil activation marker) and immunoglobulin genes that decrease over time compared to patients with late onset delirium (Fig. 4B). *In silico* deconvolution of PBMC subtype with CIBERSORTx analysis (Fig. 4C) followed by ANOVA revealed significant main effects for delirium onset in naïve CD4 and CD8 memory T cells (*p* = 0.044 and 0.048, respectively) and main effects approaching significance in plasmocytoid dendritic cells. Main effects for timepoint were observed in classical monocytes *(p* = 0.045) and CD8 memory T cells (*p* = 0.024). Post-hoc tests further revealed a trend towards significant decrease in classical monocytes over time only in patients with late delirium (Tukey’s HSD, *p* = 0.099).

**Figure 4.**
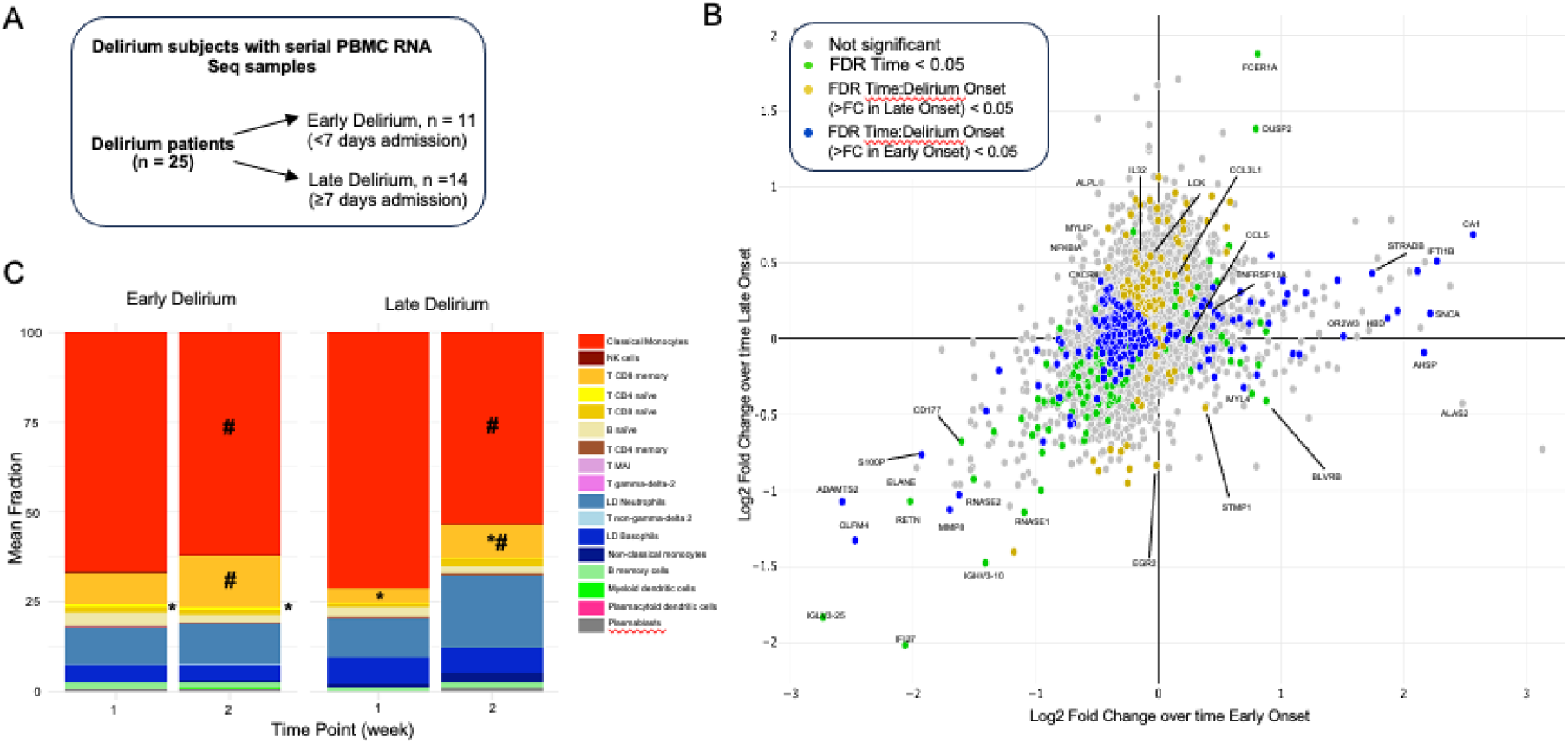
Generalized linear model (GLM) of PBMC gene expression in serial samples in patients with delirium. **(A)** Schematic of delirium patients included in the analysis. **(B)** Scatter plot comparing longitudinal gene expression changes between early (< 7 days, n = 11) or late (≥ 7 days, n = 14) delirium of paired PBMC RNA seq samples during weeks 1 and 2 of hospitalization. Log_2_ fold change in early and late delirium expression is represented on the *x* and *y*Daxis, respectively. Genes equally affected by each drug lie along the line of identity (dashed). Fold change and statistical analysis of longitudinal differential gene expression were calculated by negative binomial general linear mixed-effects model. Genes in green show significant (FDRD<D0.05) overall change in expression over time; those in blue/yellow show significantly differential change in expression over time between delirium onset based on significant (FDRD<D0.05) interaction term *delirium onset*D*×*D*time.* **(C)** *in silico* deconvolution of PBMC fractions using CIBERSORTx demonstrates increased activated CD4 naive T cells in early delirium and decreased CD8 memory T cells in late delirium. Classical monocytes and CD8 memory T cells were also significantly different over time. *Indicates main effect for Delirium onset, *p <* 0.05) # indicates main effect for time (*p <* 0.05). LD: low density; MAI: mucosal associated invariant.

### Corticosteroid treatment modulates delirium associated gene expression profiles

Early administration of dexamethasone is a key treatment for COVID19 in patients requiring supplemental oxygen. Methylprednisolone was also given at our center for severe COVID-19 infection prior to clinical trials using dexamethasone. Dexamethasone and other corticosteroids modulate immune responses and are a major risk factor for the development of delirium although the underlying mechanisms are not clear ^9,15^. To gain additional insights into mechanisms associated with delirium following IV corticosteroid treatment, we further examined changes in gene expression stratified by corticosteroid treatment and delirium (patients receiving least 3 days of IV corticosteroid treatment and developing delirium following initiation of treatment; Fig. 5A). In patients receiving corticosteroids, severity-adjusted analysis revealed 2039 genes were differentially regulated over time between delirium and control patients (FDR < 0.05, Supplementary Data File). Patients not receiving corticosteroid treatment demonstrated 622 genes significantly altered over time between delirium and control (Supplementary Data File). There was overlap in 238 significantly regulated genes in patients with delirium regardless of corticosteroid treatment. In this overlapping gene set, GO:BP analysis implicated immune and inflammatory gene pathways including platelet activation, TNF production, and leukocyte migration (Fig. 5B). Two genes demonstrated elevated expression over time in patients with delirium compared to non-delirium patients regardless of receiving corticosteroids or not: dual phosphatase specificity (*DUSP*)-2 and Kruppel-like factor (*KLF*)-10 (Fig. 5C). DUSP2 and KLF10 have previously been implicated in immune functions, aging, and metabolic dysfunction. Interestingly, the trajectory of expression of platelet activation (i.e. *ITGB3*, *PF4*) and immune-related (*NFKB1*, *TGFB1*) genes exhibited divergent trajectories in delirium patients dependent on corticosteroid administration (Supplementary Figure 4). Genes upregulated in delirium patients receiving steroids, but not control patients receiving steroids may be particularly relevant to the pathophysiology of corticosteroid-related delirium in COVID-19, and included complement C1 q B chain (*C1QB*), *CCL5*, *CXCR4*, *IL32*, and numerous nuclear-encoded oxidative phosphorylation genes such as NADH:ubiquinone oxioreductase (*NDUFA*)-*4* and cytochrome c oxidase (*COX*)-*4I1* (Fig. 5D).

**Figure 5.**
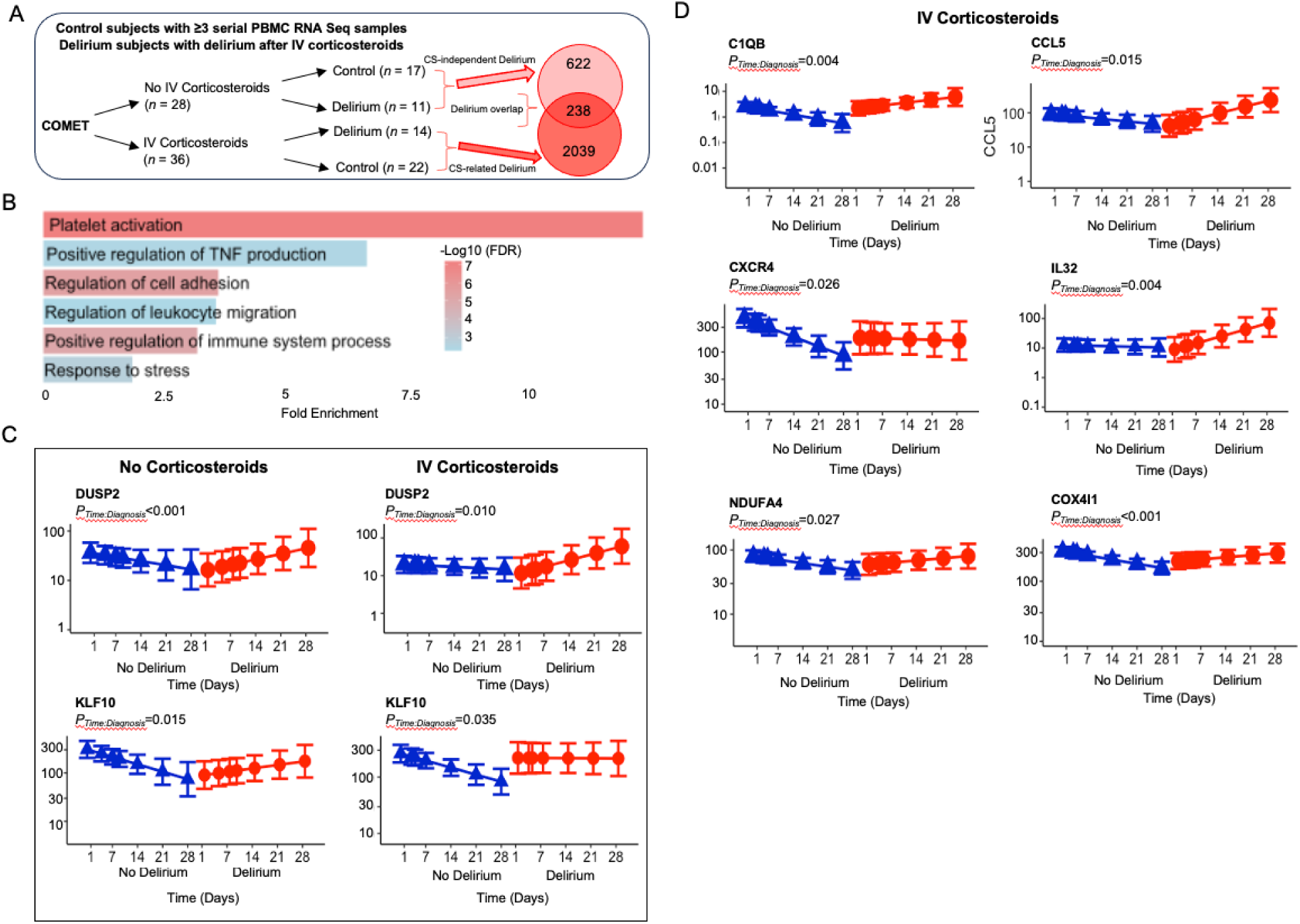
Generalized liner mixed model (GLMM) demonstrating interaction of corticosteroid administration in serial samples. **(A)** Schematic of patients and samples included in the GLMM analysis and Venn diagram demonstrating overlapping genes regulated by the interaction of *delirium x time* in patients not treated with IV corticosteroids (top) and those treated with IV corticosteroids (bottom). **(B)** GO:BP analysis of overlapping genes found to be significant for *diagnosis x time* interaction in both patients treated with and without IV corticosteroids in severity adjusted models. **(C)** Plots for notable genes similarly regulated by delirium regardless of corticosteroid treatment **(D)** Plots for selected genes specifically upregulated in patients treated with corticosteroids and developing delirium (red) compared to those not developing delirium (blue). Colored lines show regression line of fitted mixed-effects model, with error bars showing 95% CIs (fixed effects).

## DISCUSSION

Our approach differs substantially from previous efforts to characterize transcriptomic responses associated with delirium in hospitalized patients. First, we analyzed RNA from serial samples of plasma in a well-characterized cohort with a known infection. Second, we performed complementary generalized linear mixed model analyses to further understand how PBMC transcriptomic trajectories differ in patients with delirium, their interaction with delirium onset and corticosteroid treatment, and genes specifically associated with delirium onset and resolution. This high-dimensional readout informs our understanding of the pathophysiology of delirium in hospitalized patients with an infectious illness. The association of delirium with dysregulated immune responses has been described, however, we extend this by highlighting differences in pathophysiology in early compared to late delirium and following corticosteroid treatment. For example, in severity adjusted analyses, PBMC transcripts classically involved in innate immune responses such as complement pathways, cytokine production, and monocyte/macrophage recruitment exhibited an increased expression trajectory over time in delirium patients (e.g. *CCL2*, *CR1L*, *C1QA*, *TARM*, and *SLC6A19)*. Genes predominantly involved in adaptive immune functions such as T-cell activation were initially elevated and then exhibited a decreased trajectory of expression over time in delirium patients and included *CCR5*, CXCR3, *IL18R1*, *CD28*, and *NFATC2*. Interestingly, our analysis also implicated novel pathways such as stress granule assembly in delirium, which may reflect a response to excessive cytokine production and an interesting pathway for further study in delirium ^16–18^. These results are consistent with the hypothesis that altered peripheral immune responses in early COVID-19 infection are important contributors to later delirium risk

Analyses of the corticosteroid-related modulation of gene expression also provided intriguing insights concerning the pathophysiology of delirium. Corticosteroid administration had a potent effect on several mononuclear cell subpopulations (Supplementary Fig. 4), and notably reduced *in silico* predicted low density neutrophil fractions after treatment, consistent with potentially beneficial acute effects to reduce inflammation in severe COVID-19 ^19–21^. Despite this, delirium patients receiving corticosteroids demonstrated persistent immune dysregulation with prolonged elevations in inflammatory mediators such as *C1qB*, *CCL5*, *IL32*, *CXCR4*, *TGFB1*, and *NFKB1* signaling genes. These markers were comparatively decreased in delirium-free patients not receiving corticosteroids, supporting the role of corticosteroid-specific immunomodulation contributing to delirium. Corticosteroid treatment also influenced the metabolic phenotype of PBMCs specifically in delirium patients, as numerous nuclear-encoded mitochondrial genes such as *NDUFA4* and *COX4I1* were upregulated over time compared to control patients. Altered metabolic phenotypes of PBMCs may also be an important factor in persistent immune dysfunction ^22^. Interestingly, GO:BP enrichment analysis pointed to a role for the modulation of platelet activation in delirium regardless of corticosteroid treatment. Amongst PBMCs, the altered expression of platelet related genes such as *PF4* and *ITGB3* may point to B-cell and dendritic cell involvement, which also express these genes ^23,24^. This may represent a promising angle for future research, as platelet count and activity has previously been associated with post-operative delirium ^25,26^. The GLMM analyses also demonstrated that *DUSP2* and *KLF10* have a robust association with delirium regardless of corticosteroid administration. These are also promising targets for future study as *DUSP2* is a critical regulator of signal transducer and activator of transcription (*STAT*)-3 and *DUSP2* activation is known to impair CD4+ helper T (Th)-17 cell differentiation and host defense responses ^27,28,29^. *KLF10* is an important target of *TGF-β* and modulates T-cell function and Th17 responses ^30,31^. Taken together, our results point pathophysiological roles for corticosteroids promoting persistent aberrant immune responses which may contribute to delirium risk over time. Further understanding how different etiologies such as infection or medications converge on common pathways such as immune-related (Th17 and TGF*-β* responses), platelet activation pathways, stress granule assembly, and/or mitochondrial pathways to cause delirium is of particular importance for future research.

Our longitudinal sequencing reveals dynamic aberrant patterns in the initial immune response accompanying delirium with an exuberant innate immune and deficient T-cell response that normalizes with the resolution of delirium. From a clinical perspective, further understanding delirium pathophysiological responses over time could significantly contribute to the identification of new targets and key times for intervention with novel anti-delirium therapeutics in patients at risk for delirium. Focusing on PBMCs as in infectious delirium is promising since inflammatory cells and signaling molecules reflect severity of illness and may in turn influence blood-brain barrier permeability and neuronal function ^14^. Our results implicate gene pathways known to be involved in delirium such as TNFα and oxidative phosphorylation as well as novel pathways including stress granule assembly, platelet activation, and mediators of TGF-β signaling (DUSP2 and KLF10).

Although this is the largest series of delirium patients with longitudinal PBMC transcriptomic data, a major limitation of this study is the modest sample size, making it difficult to disentangle the contribution of other important delirium-related factors such as age and medications received. Refining and validating our prediction model in an independent cohort and incorporating additional demographic and clinical information will greatly improve the clinical utility of a predictive signal. Moreover, our study used a mix of delirium screening tools to capture episodes of delirium throughout the duration of the hospitalization regardless of patient location in the hospital. Future incorporation of additional standardized assessment measures or neurological examinations could offer more detailed insights into patients’ conditions and enable more accurate subclassification of delirium and outcome predictions. Another limitation regarding interpretation of the longitudinal results is that patients with less severe COVID-19 were discharged sooner and had fewer longitudinal samples available for study, introducing some degree of bias. Distinguishing delirium pathophysiology in patients with longer hospitalizations may be more clinically relevant to future predictive studies.

In conclusion, this study provides insights from analysis of PBMCs regarding the mechanisms driving delirium development in hospitalized patients with COVID-19. Identifying the dynamic patterns of regulation of PBMC transcripts accompanying delirium may facilitate the development of novel pharmacological interventions, and our results suggests that further investigation of targeting deficient Th17 signaling or hyperactive TGFβ signaling driven by DUSP2 and KLF10 may lead to promising anti-delirium therapeutics.

## Data Availability

All data produced in the present work are contained in the manuscript and source data is available online.

https://doi.org/10.5061/dryad.rv15dv4gw

## List of Supplementary Materials

Fig. S1: Additional delirium cohort information

Fig. S2: Additional mixed effects gene model of serial PBMC sequencing

Fig. S3: Additional mixed effects gene models of serial PBMC sequencing in patients treated with or without steroids

Fig. S4: CIBERSORTx deconvolution for the effects of IV corticosteroid treatment on PBMC cell fractions

Supplementary Data File

## Funding

National Institutes of Health grant U19AI077439 (DJE, CSC, PGW, MFK)

National Institutes of Health grant R01NS125693 (MRW, SJP)

Chan Zuckerberg Foundation 2019-202665 (WE, DJE)

Genentech (COMET Plus, TSK-020586 to UCSF COMET consortium)

## Author contributions

Conceptualization: SCL, SJP, MRW, NeSS

Methodology: NT, RS, AB, EM, AS, TBC, NiSS

Investigation: NT, RS, AB, EM, AS, TBC, NiSS

Visualization: NT, RS, NeSS

Funding acquisition: WE, DJE, CSC, PGW, MFK, MRW, SJP Project administration: WE, DJE, CMH

Supervision: WE, CRL, MFK, PGW, NeSS

Writing – original draft: RS, AB, NeSS

Writing – review & editing: SCL, NT, RS, CMH, CSC, DJE, TO, MS, AF, VCD, JCN, SJP, MRW

## Materials and Correspondence

RNA-sequencing processed data and metadata can be accessed at Dryad data repository.

## Supplementary Figure Captions

**Supplementary Figure 1: Additional delirium cohort information.** Subjects included in the longitudinal delirium analyses and receiving intravenous corticosteroids (IVCS) are denoted by filled in circles. Adjacent bar plot represents hospital days prior to (blue), during (red), and following the resolution of delirium (green).

**Supplementary Figure 2: Additional mixed effects gene model of serial PBMC sequencing.** Plots for selected genes with colored points showing regression line of fitted mixed-effects model, with error bars showing 95% CIs (fixed effects)

**Supplementary Figure 3. Additional GLMM gene model plots separated by corticosteroid treatment and delirium.** GLM demonstrating interaction of corticosteroid administration in serial samples (A) Plots for platelet activation genes and (B) inflammation-related genes significantly regulated by delirium in patient not receiving and receiving corticosteroid treatment. Patients developing delirium (red) are compared to those not developing delirium (blue). Colored lines show regression line of fitted mixed-effects model, with error bars showing 95% CIs (fixed effects).

**Supplementary Figure 4: CIBERSORTx deconvolution for the effects of IV corticosteroid treatment on PBMC cell fractions.** *in silico* deconvolution of PBMC fractions using CIBERSORTx demonstrates increased low density (LD) neutrophils at admission in patients receiving corticosteroids decreased following steroid treatment. LD basophils were increased following steroid treatment. *, *p <* 0.05; ***, *p <* 0.001 compared to IV corticosteroid admission (week 0); #, *p <* 0.05; ##, *p <* 0.01 compared to No corticosteroid admission (week 0) by Tukey’s HSD post-hoc test. There were main effects by ANOVA for steroid treatment (*p* = 0.018), time point (*p* = 0.030), and *steroid:time point* (*p* = 0.032) for LD neutrophils. ANOVA also revealed main effects for steroid treatment were present for NK cells *(p* = 0.027), CD8 Memory T cells (*p* = 0.003), Non-V delta 2 gamma delta T cells (*p* = 0.011), and plasmacytoid dendritic cells (*p =* 0.013). LD: low density; MAI: mucosal associated invariant.

