## Supplementary figures and images for "Peripheral blood mononuclear cell transcriptomic trajectories reveal dynamic regulation of inflammatory actors in delirium"

### Supplementary Figures 1-4

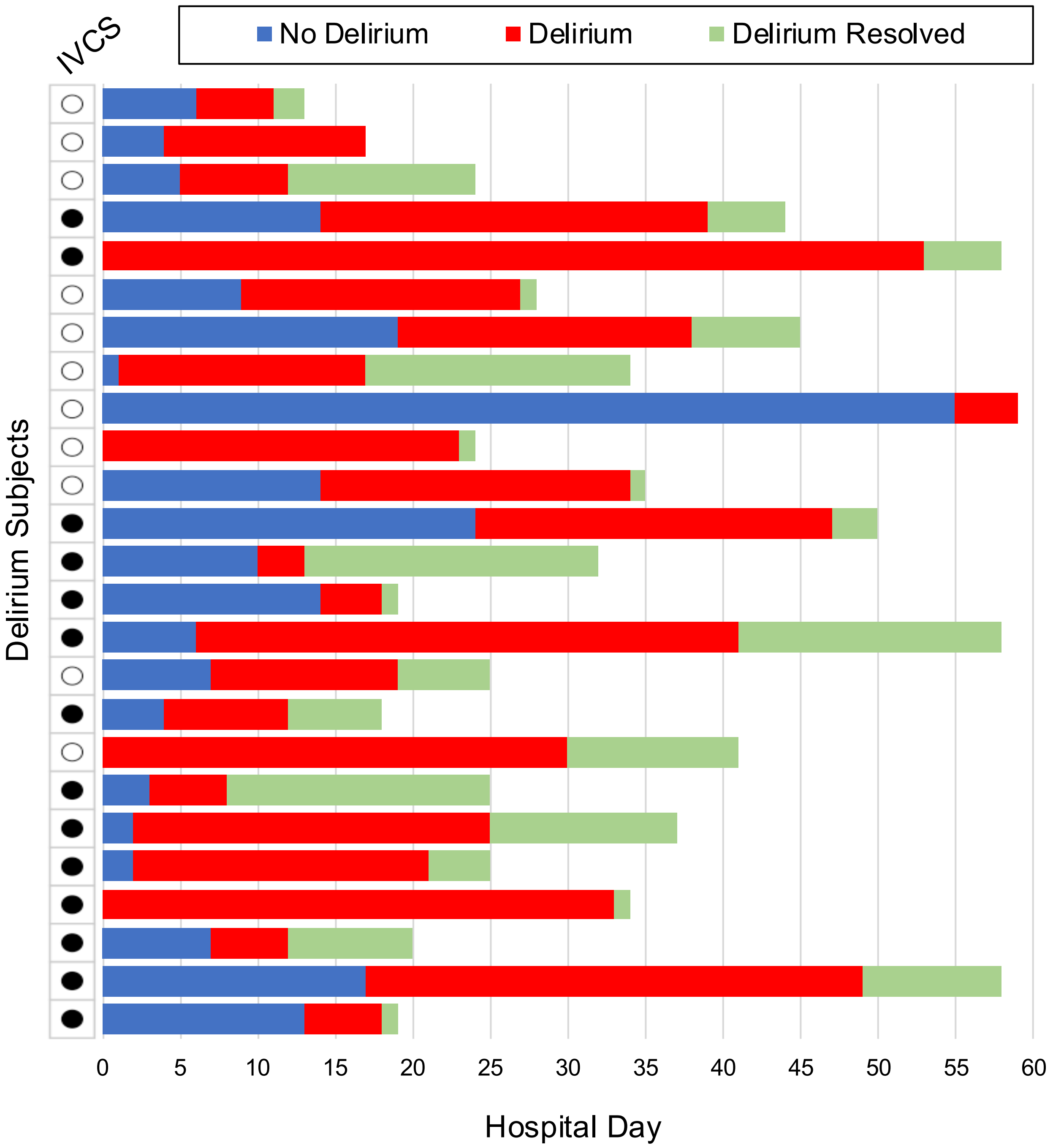

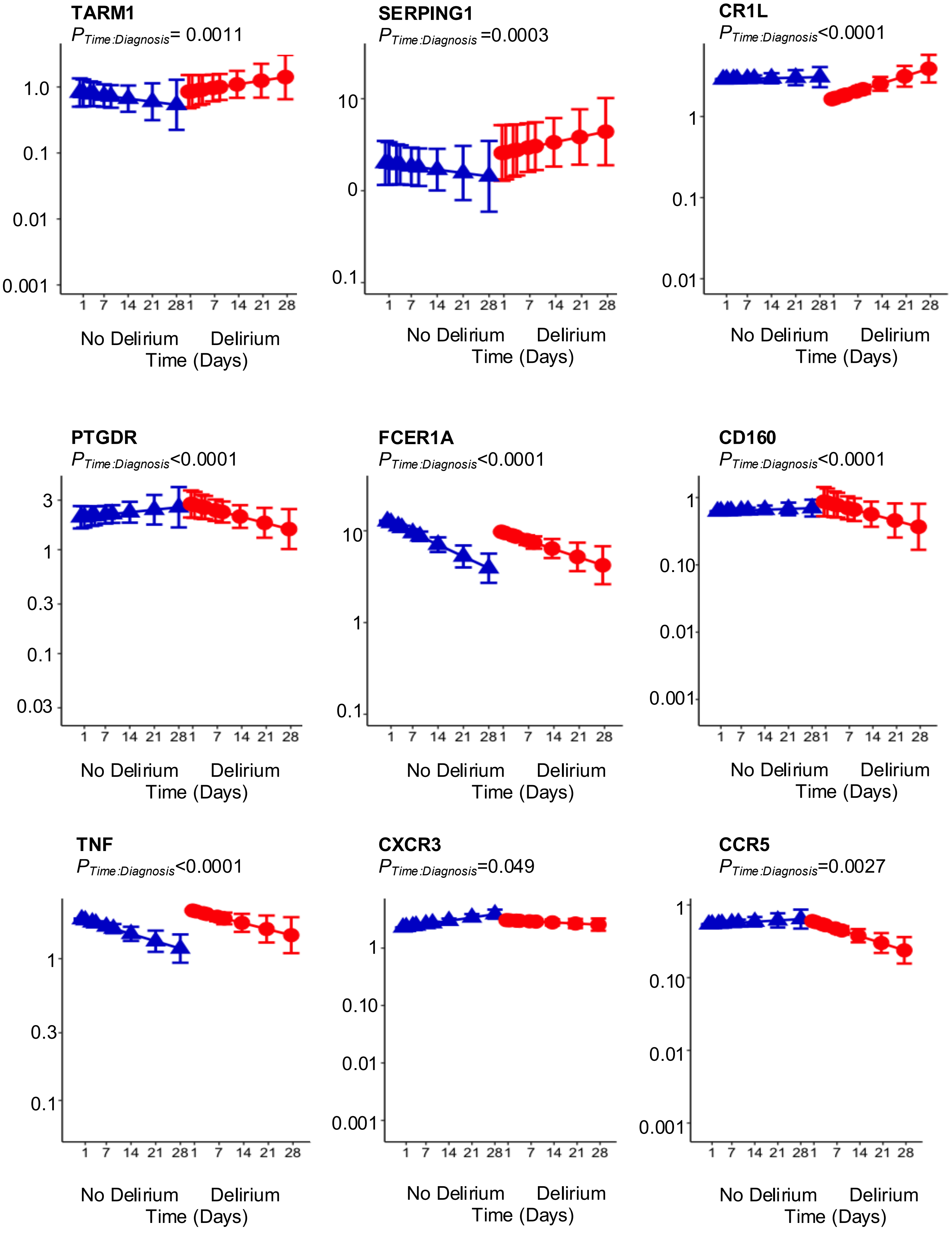

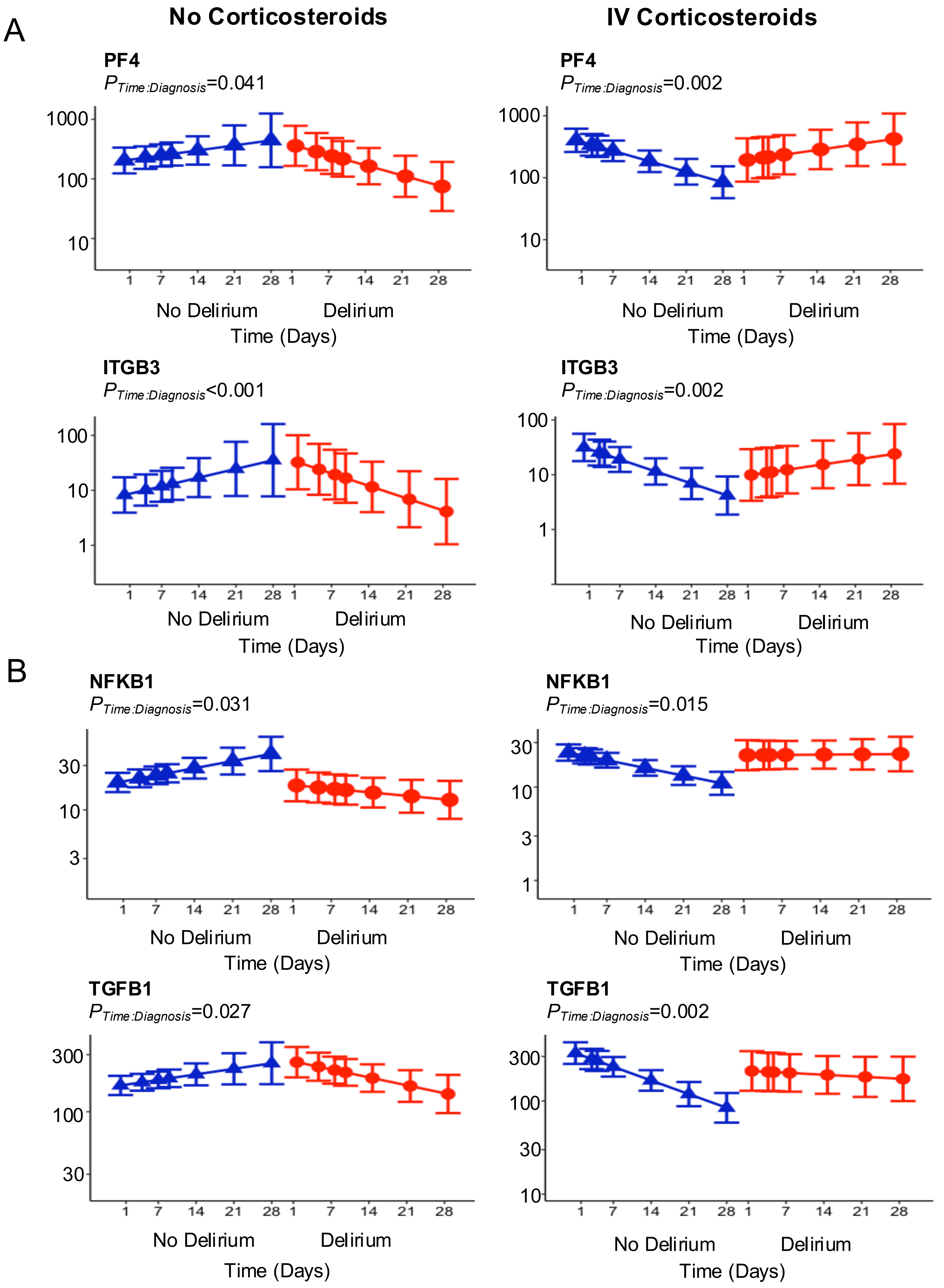

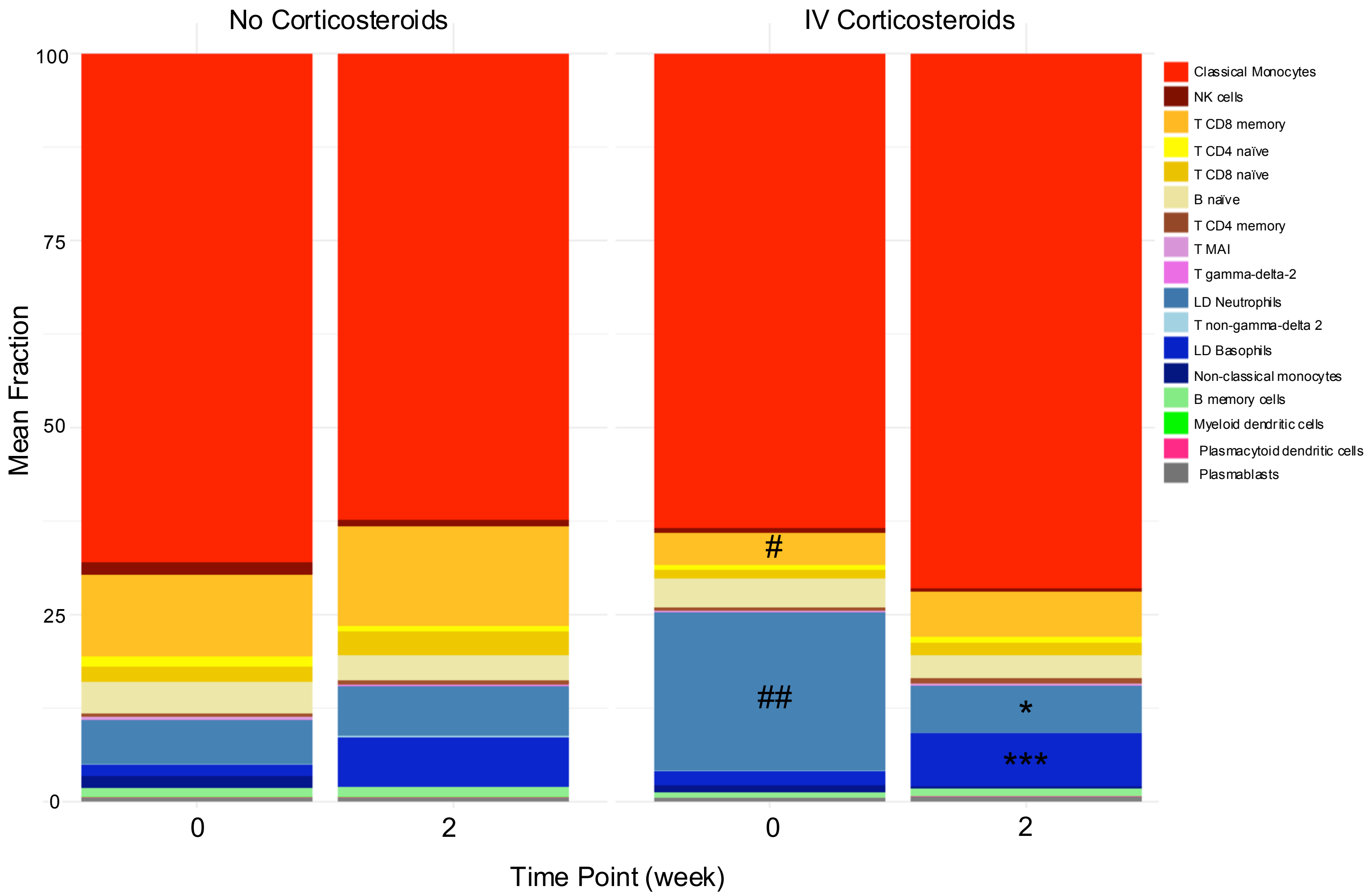
